# Feasibility study of a randomised controlled trial of pre and postoperative nutritional supplementation in major lung surgery

**DOI:** 10.1101/2021.09.08.21263295

**Authors:** Amy Kerr, Sebastian T. Lugg, Salma Kadiri, Amelia Swift, Nikolaos Efstathiou, Krishna Kholia, Venessa Rogers, Hazem Fallouh, Richard Steyn, Ehab Bishay, Maninder Kalkat, Babu Naidu

## Abstract

**Objectives:** Malnutrition and weight loss are important risk factors for complications after lung surgery. However, it is uncertain whether modifying or optimising perioperative nutritional state with oral supplements results in a reduction in malnutrition, complications, and quality of life.

**Design:** A randomised, open label, controlled feasibility study was conducted to assess the feasibility of carrying out a large multicentre randomised trial of nutritional intervention. The intervention consisted of preoperative carbohydrate-loading drinks (4x 200mls evening before surgery and 2x 200mls the morning of surgery), and early postoperative nutritional protein supplement drinks twice a day for 2 weeks following surgery compared to the control group receiving an equivalent volume of water. Qualitative interviews were conducted with participants to help determine the acceptance of the study.

**Setting:** Single adult thoracic centre in the UK.

**Participants:** All patients admitted for major lung surgery. Participants were included if were able to take nutritional drinks prior to surgery and were able to give written informed consent. Patients were excluded if they were likely unable to comply with completion of the study questionnaires, they had a body mass index (BMI) < 18.5 kg/m^2^, were receiving parenteral nutrition or known pregnancy.

**Results:** All patients presenting for major lung surgery were screened over a 6-month period, with 163 patients screened, 99 excluded and 64 (41%) patients randomised. Feasibility criteria were met and the study completed recruitment 5 months ahead of target. The 2 groups were well balanced, and tools used to measure outcomes were robust. 97% of patients were compliant with pre-surgery nutritional drinks and 89% of the questionnaires at 3 months were returned fully completed. The qualitative interviews demonstrated that the trial and the intervention were acceptable to patients. Patients felt the questionnaires used captured their experience of recovery from surgery well.

**Conclusion:** A large multicentre randomised controlled trial of nutritional intervention in major lung surgery is feasible. It is possible to randomise eligible patients and follow up with high fidelity. A pre-op carbohydrate-loading and post-surgery supplementation is highly acceptable to patients with good compliance to both intervention and trial measures. A large multi-centre clinical trial is required to test clinical efficacy in improving outcomes after surgery.

**Trial registration number:** ISRCTN16535341

**Strengths and limitation of this study:**

- This randomised, feasibility study had pre-planned feasibility to assess whether a larger randomised trial would be feasible.
- The study included a large regional thoracic surgical centre and cohort of patients undergoing major lung surgery predominantly for cancer, which would be representative of full trial national recruitment.
- The study was not designed and powered to be large enough to provide conclusive evidence to support the use of nutritional intervention in major lung surgery, but provided evidence that a larger, substantive randomised controlled trial is feasible.

## INTRODUCTION

In the UK patients undergo major lung surgery (MLS), including over 6,500 resections per year for treatment of lung cancer.^1^ Postoperative pulmonary complications (PPC) occur in 13% of all MLS patients. Once a PPC has developed, there is an increase in mortality (1 to 12%), intensive treatment unit (ITU) admission rate (2 to 28%), length of hospital stay (6 to 13 days) and 30-day hospital readmission (12 to 21%).^2^ Poor nutritional state is a major independent risk factor for death and complications after MLS.^3^ Two thirds of all patients undergoing MLS are malnourished before or are at risk of becoming so after surgery.^4,5^

Malnutrition and a loss of muscle mass are frequent in cancer patients and are associated with poor clinical outcomes.^6^ Oral nutritional supplement drinks (ONS) are recommended in cancer patients who are malnourished or at risk of malnutrition,^6^ and as part chronic obstructive pulmonary disease (COPD) rehabilitation programmes; COPD is common in patients with lung cancer.^7^ The use of ONS in COPD patients is associated with weight gain, improved patient quality of life and respiratory muscle function particularly in the undernourished.^8^ A meta-analysis of nutritional interventions including ONS in cancer patients also showed improvements in both weight gain and quality of life.^9^

There are two distinct types of nutritional intervention as discussed in the Enhanced Recovery after Lung Surgery guidelines;^10^ carbohydrate loading before (CHO) surgery and perioperative protein nutrition supplementation via ONS or enteral nutrition. Surgery induces a catabolic response with release of stress hormones and inflammatory mediators, with resultant cellular dysfunction and insulin resistance.^11^ The hyperglycaemia and reduced insulin-stimulated glucose uptake in skeletal muscle and adipose tissue persists for days following surgery and is associated with increased morbidity, mortality and length of hospital stay.^11,12^ In other types of major surgery, preoperative CHO loading has been demonstrated to ameliorate the physiological hit of surgery to metabolic, muscle and lung function and also improved length of stay.^13,14^ CHO loading has also been shown to significantly reduce patient symptom burden.^15,16^ in major abdominal surgery, routine pre and/or postoperative ONS reduced postoperative weight loss, improved nutritional status and muscle strength and may reduce complication rates.^17,18^

The most recent thoracic surgery specific guidelines cite the evidence for nutritional intervention as being of moderate level.^10^ The European nutritional guidance in surgery ranked evidence only ‘as good practice’ rather than of high level (A or B).^19^ A national survey of all 38 UK thoracic surgery units found that almost all patients undergoing lung surgery are not routinely offered either CHO or ONS.^20^ Despite CHO loading recommended is the Enhanced Recover After Surgery (ERAS) guidelines, evidence is limited in MLS, and only few reports of its use in practice.^21^ One small study in MLS of nutritional supplementation (n=58), where patients were randomised to receive 10 days of preoperative immune enhancing nutrition or normal diet.^22^ The with nutritional intervention had reduced plasm albumin drop and a reduction in PPC incidence, though they classified air leak as a PPC despite this being a minor surgical complication. It was also under powered to detect any difference in clinical outcomes.^22^ Thus, the gap in direct evidence prevents strong guidance for ONS and CHO loading in MLS. It’s clear that the type and magnitude of surgery are important in the efficacy of CHO loading,^13,14^ and so it is important to conduct an independent study in patients undergoing MLS. Through nutritional supplementation one might be able to optimise recovery after surgery and preventing complications.^23^

The aim was to conduct a single centre mixed method open label randomised controlled trial (RCT) to assess the feasibility of carrying out a large multicentre RCT in MLS patients (ISRCTN: 16535341). We compared a nutritional intervention regime of preoperative CHO drinks and postoperative ONS to a control group receiving an equivalent volume of water.

## METHODS

We conducted a randomised controlled feasibility trial of carbohydrate loading drinks pre-surgery and high energy, high protein drinks post-surgery compared to the equivalent volume in water in enhancing recovery after major lung surgery. Recruitment took place over 6 months in an adult thoracic centre at Heartlands Hospital, Birmingham.

### Population and inclusion/exclusion criteria

Eligible participants were all adults aged over 18 undergoing elective major lung surgery (MLS). MLS was defined as any patient having part of the lung removed for primary or secondary cancer with curative intent or benign lung conditions. Additional eligibility included participants able to consume nutritional drinks prior to surgery and were able to give written informed consent. Participants were excluded if they were likely unable to comply with completion of the study questionnaires, they had a body mass index (BMI) < 18.5 kg/m^2^, were receiving parenteral nutrition or known pregnancy.

### Study conduct

Patients listed for MLS were identified and screened for eligibility at clinics prior to surgery. When a patient was screened but not eligible for the trial or did not consent for randomisation, a record of the case was been recorded on a detailed screening log. This data informed recruitment targets and help with sample size in the definitive trial and answered the feasibility outcome questions. Participants were provided with a Patient Information Sheet about the study, including details of the treatment procedures and trial data collection. Written valid informed consent was obtained from each of the study participants under unhurried circumstances. Participants were informed of the aims, methods, any conflicts of interests, benefits and risks of participating in line with International Conference on Harmonisation (ICH) Good Clinical Practice (GCP). Re-confirming consent was sought at every study contact and participants could withdraw consent at any time without any reprisal. After written informed consent, the patients were randomised before surgery to either a nutritional intervention or water. Participants were individually randomised into the study in an equal 1:1 ratio; randomisation was conducted using a web-based randomisation system. Patients were stratified by diagnosis (cancer or benign) and type of surgery (video-assisted thoracoscopic surgery (VATS) or open thoracotomy) this stratification and online randomisation service assisted to reduce allocation bias.

### Interventions

The nutritional intervention was defined as follows. In the preoperative period, the evening before surgery patients consumed 4x200ml of carbohydrate-loading supplement. On the morning of surgery patients consumed 2x200ml of carbohydrate-loading supplement and if the surgery was scheduled for the afternoon, 200 mls of carbohydrate loading drinks was given every two hours up until 2 hours before surgery. This has proven to be the most effective regime of carbohydrate-loading loading in terms of insulin resistance^24^ (Nutricia preOp, per 100ml: 50 kcal, 12.6g carbohydrates, 0g protein, 290 mOsm/kg, pH 5.0). In the postoperative period, patients were given 125ml polymeric nutritional supplement drink twice daily from the period immediately after their operation until discharge, continuing at home up until 14 days after surgery (Fortisip Compact Protein, per 100ml: 240kcal, 24.4g carbohydrate, 14.4g protein, 900 mOsm/kg, pH 6.6).

The control group was provided with the same quantity of water in bottles to take home, thus any benefit from the intervention will not be due to preventing dehydration. All other aspects of the patient care was as per usual care in the both groups. All patients received standard patient information based on current national guidelines. Free fluids were permitted immediately after surgery and a light diet as tolerated by the patient. Standardised nausea and vomiting prophylaxis and laxatives prescribed. Analgesic technique is based on patients’ preference and discussion with the anaesthetists. Both the intervention and control groups were managed daily by a specialised thoracic team and all received a daily physiotherapy programme from postoperative day (POD) 1 onwards.

### Feasibility outcomes

The following outcomes and targets provided the basis for interpreting the results of the study and determining whether it is feasible to proceed to the substantive study. The primary outcome was patient recruitment rate: It is estimated that 300 eligible patients a year will undergo MLS a recruitment rate of 5 a month for 12 months, i.e. 60 patients.

Secondary objectives included: (1) Reasons for failure to recruit. (2) Is the randomisation process of patients easy to use and efficient? This was ascertained by the speed in which patients can be randomised and whether important prognostic data can be collected pre-operatively. (3) What is the compliance rate of the intervention and contamination rate of the control group? Data was gathered by questionnaires and interviews; we expected to have a compliance of 50% of prescribed carbohydrate drinks and ONS taken as scheduled. (4) Are the data collection processes during patient’s hospital stay robust? We expected completeness of important peri-operative data to be over 90%. (5) What is follow-up rate of patients at 3 months? To be viable as a primary outcome, we expected to achieve a response rate of 80%. (6) What are the reasons for loss of follow-up if any? We should be able to capture 100% of mortality data. (7) Which questionnaire best reflects patient experience? We envisaged from the patient interviews in both patients getting the intervention and those not we would be able to discern if one of the questionnaires was better than other. (8) What is the variability and distribution of quality-of-life questionnaires measured up to 3 months after surgery? This would help us ascertain an appropriate sample size for any possible full study.

### Clinician and participant-reported outcome measures

Data was collected using a Case Report Form (CRF), this included demographic information, co-morbidities, PPC was defined by the Melbourne Group Scale (MGS) this tool was selected as it has been shown to outperform other scoring tools in the recognition of PPC in lung resection,^25^ in addition to this hospital re-admission rate within 30 days of discharge was also collected. A nutritional assessment using Malnutrition Universal Screening Tool (MUST)^26^ was completed pre-operative and 3 months postoperatively, the MUST tool was used as it is fast and simple to use with a fair-good to excellent concurrent validity.^27^ Handgrip strength was used, which is a measure of muscle strength which can be used in the assessment of sarcopenia. Guidelines recommend measuring handgrip strength in thoracic patients due to their increased risk of malnutrition and sarcopenia.^28^ Handgrip strength is a simple measure that can be used in hospital and community settings, and it has been shown to be predictive of survival in advanced cancer patients.^29^ Peak expiratory flow rate was used to assess pulmonary function, as demonstrated in other studies.^30^

Recently there has been an increase in the use of patient reported outcome measures in trials. In this study, the feasibility of a number of questionnaires was tested at different stages throughout the patients’ surgical journey. Systematic reviews of postoperative recovery outcomes measurements appraised the Quality of Recovery Score-40 (QoR-40) as being a good measure of early recovery after surgery and suggested as a valuable endpoint in clinical research,^31,32^ this score has been validated in many specialities of surgery in several countries. The QoR-40 tool has 40 questions which assess 5 dimensions: physical comfort (12 items), emotional state (9 items), physical independence (5 items), psychological support (7 items) and pain (7 items) all relevant in thoracic surgery. A generic health related quality of life tool (EQ-5D-5L) was selected in this trial as it is a widely used tool and easy to use. The EQ-5D-5L has 5 dimensions of health which assess mobility, self-care, usual activities, pain/discomfort, and anxiety/depression on a 5-level classification score. The tool has been developed from the EQ-5D-3L to be more sensitive when assessing quality of life.^33^ In addition, a Visual Analogue Score (VAS) looking at patient well-being was created, these were based on questions that have been used in other RCTs assessing carbohydrate loading drinks and nutritional support in other specialties.^34,35^

Throughout the trial adverse events were collected and recorded to assess any relation between adverse events and the trial intervention. However, patients undergoing thoracic surgery have a 13% risk of developing PPCs which impact on ITU admission, length of stay, readmission and mortality.^2^ PPCs, acute complications, length of stay, readmission, ITU admission and mortality data was collected in this study therefore they were not reported as adverse events and serious adverse events. Any other AE which occurred during the duration of the patient’s involvement in the study was recorded and reported in accordance with ICH GCP guidelines.

### Statistics

We expected to recruit 60 patients over a 12-month recruitment period, 5 patients a month, depending on the number found to be eligible. Feasibility outcomes were considered with simple summary statistics including percentages. Patient reported outcomes measures were analysed with mean and mean differences and 95% CIs. Continuous data was analysed using unpaired t-test with Welch’s correction for parametric data, and Man Whitney U for non-parametric data. Categorical data was analysed using Chi squared and Fisher’s Exact Test. P value <0.05 was considered to be statistically significant. SPSS Version 27 and Prism Version 8 were used for statistical analysis. Participants were considered in the group if they were randomised which was regardless of the participants compliance (intention-to-treat).

### Qualitative assessment and survey of practice

Semi-structured telephone patient interviews were undertaken at 3-4 weeks post discharge. This time point was selected as it was 1-2 weeks after patients had finished the trial interventions and after completion of 3-week patient reported outcome questionnaires. All patients consenting to take part in the trial were eligible for interview and were selected using maximum variation sampling by age, sex, admitting diagnosis and surgical procedure. This approached was used to understand how different groups of people viewed the trial. Interviews were conducted until saturation was achieved.

An interview framework was developed using evidence from the use of nutritional intervention with other patient groups. Interviews explored the pre and post-surgical experiences of patients and the trial interventions, questionnaires and compliance issues. All interviews were digitally recorded, transcribed and coded using NVivo. Thematic analysis was used to identify the main acceptability issues for patients, and key barriers and facilitators in the use of the interventions.

### Patient involvement

Patient and public collaboration was sought from the UK thoracic surgery patient involvement group (RESOLVE). This was for creating the trial protocol, assessing, and defining outcome measures, data collection tools and writing patient information sheets and consent forms.

### Ethics

Sponsorship was gained from the University Hospitals Birmingham NHS Foundation Trust. Ethical approval was granted from Wales REC 7 research ethical committee REC reference 16/WA/0254, followed by HRA approval. The study was then registered with ISRCTN number: 16535341. The study was funded by Birmingham Health Partnership and Nutricia provided the funding for the nutritional intervention, which enabled the trial eligible for portfolio support from the Clinical Research Network in England.

## RESULTS

### Participants and follow-up

Adult patients undergoing elective MLS at a regional thoracic surgery unit were approached between September 2016 and May 2017 as the study completed 5 months ahead of target. One hundred and sixty-three patients were screened for eligibility (**figure 1**). Of those, only 5 patients were initially deemed to not meet the exclusion criteria. Of the remaining patients, the most common reason for not including patients was that they were not approached due to reaching the recruitment quota for the week. Other reasons included patients not wanting to participate, not willing to take nutritional drinks, and not willing to complete study questionnaires. Sixty-four patients were randomised. Of those, 33 patients were allocated to receive nutritional drink and 31 patients received water. Of the patients randomised, 1 patient in the nutritional group and 2 patients in the control group did not receive allocated intervention. Baseline details of the randomised participants are in **table 1**. The randomisation process provided appropriate balance for the balancing factors; the median age of those undergoing major lung surgery was 70 (IQR 60 – 74.5), with the vast majority having lung resection for cancer (n=57; 93%) and VATS approach in 37 (61%) patients. At 3 months post-randomisation, 59/61 (97%) patients were followed up, and 54/61 (89%) of questionnaire booklets were returned.

**Figure 1.**
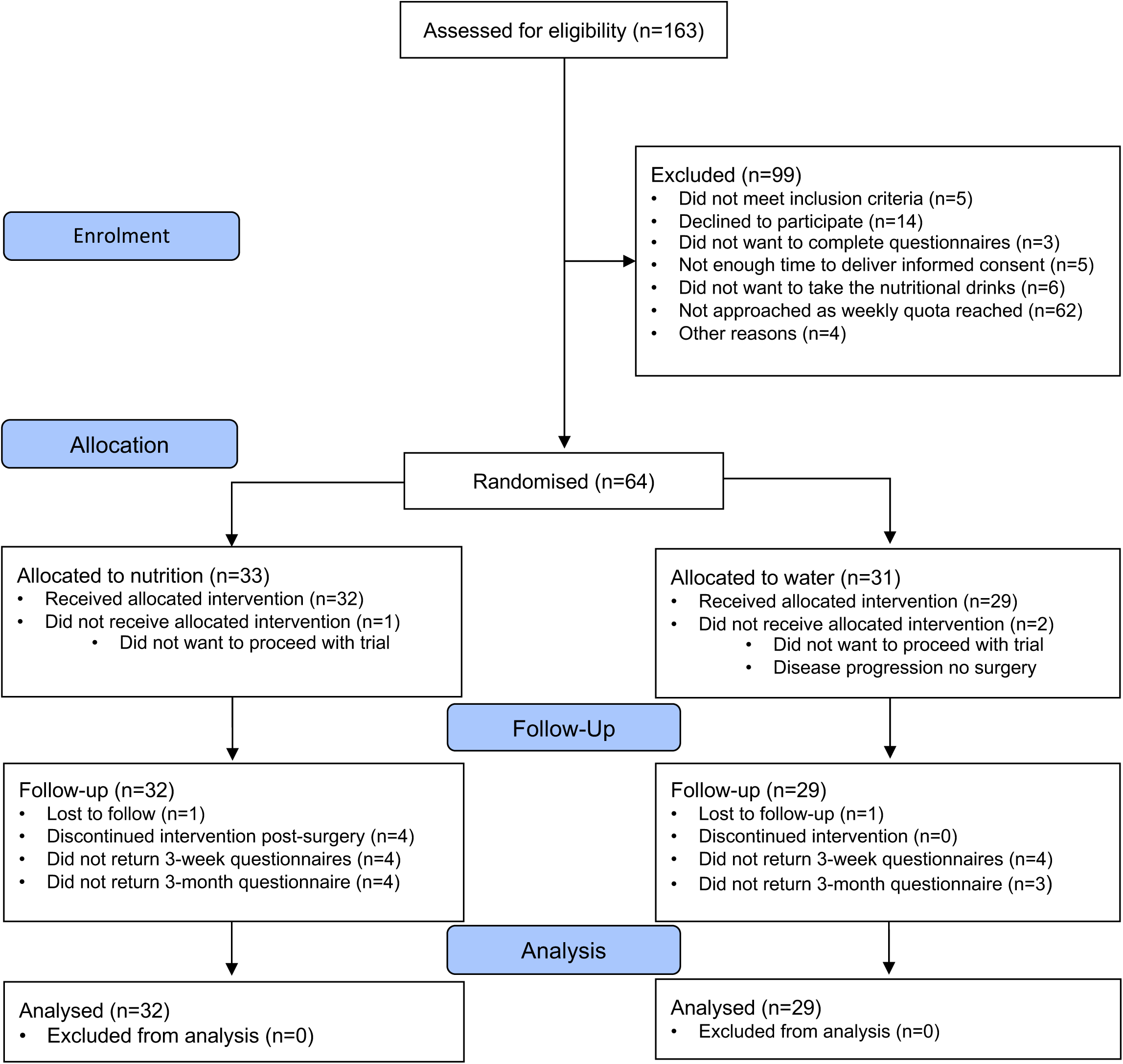
Flow of Participants during the trial.

**Table 1:** Baseline characteristics of patients

|  | **Nutrition Group**<br>**(n=32)** | **Control Group**<br>**(n=29)** | **p value** |
| --- | --- | --- | --- |
| Age years, median (IQR) | 69.5 (58.3-74) | 71 (61.5-76) | 0.355 |
| Gender no. (%) |  |  |  |
| Female | 14 (43.8) | 17 (58.6) | 0.309 |
| Male | 18 (56.6) | 12 (41.4) |  |
| BMI kg/m<sup>2</sup>, mean (SD) | 26.5 (23.4-30.1) | 27.1 (24.6-31) | 0.561 |
| ASA physical status no. (%) |  |  |  |
| 1. Normal healthy | 1 (3.1) | 0 | 0.599 |
| 2. Mild systemic disease | 16 (50) | 17 (58.6) |  |
| 3. Severe systemic disease | 15 (46.9) | 11 (37.9) |  |
| 4. Severe systemic disease with threat to life | 0 | 1 (3.4) |  |
| ECOG performance status no. (%) |  |  |  |
| 0. Normal activity | 21 (65.6) | 17 (58.6) | 0.687 |
| 1. Symptomatic but nearly fully ambulatory | 11 (34.4) | 11 (37.9) |  |
| 2. Symptomatic but ambulatory >50% of the day | 0 | 1 (3.4) |  |
| Dyspnoea |  |  |  |
| 0. No dyspnoea | 20 (62.5) | 16 (55.2) | 0.838 |
| 1. Slight dyspnoea (hurrying or walking up hill) | 9 (28.1) | 11 (37.9) |  |
| 2. Moderate (walks slower than people same age) | 2 (6.3) | 1 (3.4) |  |
| 3. Moderately severe (has to stop whilst walking) | 1 (3.1) | 1 (3.4) |  |
| Previous medical history no. (%) |  |  |  |
| Cancer | 29 (90.6) | 28 (96.6) | 0.614 |
| COPD | 5 (15.6) | 11 (37.9) | 0.079 |
| Ischaemic heart disease | 6 (18.8) | 2 (6.9) | 0.260 |
| Congestive cardiac failure | 1 (3.1) | 1 (3.4) | 1.000 |
| Hypertension | 10 (31.3) | 11 (37.9) | 0.602 |
| Diabetes | 3 (9.4) | 7 (24.1) | 0.170 |
| Renal disease | 2 (6.3) | 1 (3.4) | 1.000 |
| Previous stroke | 1 (3.1) | 3 (10.3) | 0.338 |
| Hypothyroidism | 6 (18.8) | 0 | **0.025** |
| Hyperthyroidism | 1 (3.1) | 0 | 1.000 |
| Other malignancy | 13 (40.6) | 14 (48.3) | 0.611 |
| Smoking no. (%) |  |  |  |
| Current | 2 (6.3) | 4 (13.8) | 0.238 |
| Ex-smoker <6 weeks | 3 (9.4) | 3 (10.3) |  |
| Ex-smoker 6 weeks to 1 year | 2 (6.3) | 5 (17.2) |  |
| Ex-smoker > 1 year | 17 (53.1) | 15 (51.7) |  |
| Never smoker | 8 (25) | 2 (6.9) |  |
| Pack years, median (IQR) | 15.5 (0-41) | 39 (19-46) | **0.029** |
| Alcohol units per week, median (IQR) | 2.75 (0-10) | 2 (0-18.20) | 0.689 |
| Unplanned weight loss in past 3-6 months (%) |  |  |  |
| < 5% | 31 (96.6) | 25 (86.2) | 0.316 |
| 5-10% | 1 (3.1) | 2 (6.9) |  |
| ≥ 10% | 0 | 2 (6.9) |  |
| MUST score baseline |  |  |  |
| 0 | 28 (87.5) | 23 (79.3) | 0.418 |
| 1 | 4 (12.5) | 4 (13.8) |  |
| 2 | 0 | 2 (6.9) |  |
| Anaesthetic no. (%) |  |  |  |
| Epidural | 5 (15.6) | 8 (27.6) | 0.304 |
| Paravertebral block | 6 (18.8) | 3 (10.3) |  |
| PCA | 3 (9.4) | 7 (24.1) |  |
| Morphine infusion | 2 (6.3) | 1 (3.4) |  |
| Paravertebral and PCA | 14 (43.8) | 10 (34.5) |  |
| Oral only | 2 (6.3) | 0 |  |
| Surgical technique no. (%) |  |  |  |
| VATS | 21 (65.5) | 16 (55.2) | 0.442 |
| Thoracotomy | 11 (34.4) | 13 (44.8) |  |
| Resection type no. (%) |  |  |  |
| Lobectomy | 22 (68.8) | 15 (51.7) | 0.407 |
| Segmentectomy | 1 (3.1) | 2 (6.9) |  |
| Wedge | 8 (25) | 11 (37.9) |  |
| Lung biopsy | 1 (3.1) | 0 |  |
| Other | 0 | 1 (3.4) |  |

BMI, body mass index; ASA, Association Society of Anaesthesiologists; ECOG, Eastern Co-operative Oncology Group; COPD, chronic obstructive pulmonary disease; MUST, Malnutrition Universal Screening Tool; PCA, patient controlled analgesia; VATS, video assisted thoracoscopic surgery.

### Nutritional Intervention

The consumption of drinks in both the intervention and control group is shown in **table 2**. All patients in the nutritional intervention group had 5 or more drinks prior to surgery with 96.9% (31/32) having 6 or more drinks. On POD1, 71.9% (23/32) of patients received both drinks, with 84.4% (27/32) receiving at least 1 drink. By POD3 68.8% (22/32) received both drinks, whilst 81.3% (26/32) received at least one drink. The median overall compliance of postoperative drinks between POD1-14 was 89% (25/28 drinks, IQR 14.5-28). Within the control group of water only, there was 100% compliance with all patients.

**Table 2:** Drinks consumed between groups

|  | **Nutrition Group**<br>**(n=32)** | **Control Group**<br>**(n=29)** | **p value** |
| --- | --- | --- | --- |
| Pre-surgery drinks no. (%) |  |  |  |
| 2 | 0 | 2 (6.9) | 0.137 |
| 4 | 0 | 1 (3.4) |  |
| 5 | 1 (3.1) | 1 (3.4) |  |
| 6 | 22 (68.8) | 22 (75.9) |  |
| 7 | 9 (28.1) | 3 (10.3) |  |
| POD 1 drinks no. (%) |  |  |  |
| 0 | 5 (15.6) | 0 | **0.002** |
| 1 | 4 (12.5) | 0 |  |
| 2 | 23 (71.9) | 29 (100) |  |
| POD 2 drinks no. (%) |  |  |  |
| 0 | 3 (9.4) | 0 | **<0.001** |
| 1 | 7 (21.9) | 0 |  |
| 2 | 9 (68.8) | 29 (100) |  |
| POD 3 drinks no. (%) |  |  |  |
| 0 | 6 (18.8) | 0 | **<0.001** |
| 1 | 4 (12.5) | 0 |  |
| 2 | 22 (68.8) | 29 (100) |  |
| POD 1-14 drink compliance, median (IQR) | 25 (14.5-28) | 28 (-) | **<0.0001** |

POD, postoperative day.

### Clinical outcomes

PPC incidence was lower in those of the nutritional intervention group compared to the control group (3/32; 9.4% vs 5/29 17.2%), though this was not statistically significant, which would be expected due to the study not being powered for this reason (p=0.460) (**table 3**). There were no differences between HDU stay, ITU admission and LOS between groups.

**Table 3:** In-hospital outcomes

|  | Nutrition Group<br>(n=32) | Control Group<br>(n=29) | p value |
| --- | --- | --- | --- |
| PPC (%) |  |  |  |
| No | 29 (90.6) | 24 (82.8) | 0.460 |
| Yes | 3 (9.4) | 5 (17.2) |  |
| HDU stay (%) |  |  |  |
| No | 31 (96.9) | 27 (93.1) | 0.600 |
| Yes | 1 (3.1) | 2 (6.9) |  |
| ITU stay (%) |  |  |  |
| No | 31 (96.9) | 28 (96.6) | 1.000 |
| Yes | 1 (3.1) | 1 (3.4) |  |
| Death (%) |  |  |  |
| No | 32 (100) | 29 (100) | N/A |
| Yes | 0 | 0 |  |
| Hospital LOS (days), median (IQR) | 4 (3-6) | 4 (3-6.5) | 0.336 |

PPC, postoperative pulmonary complication; HDU, high dependency unit; ITU, intensive therapy unit; LOS, length of stay.

**Table 3:** Clinical outcomes

|  | Nutrition Group<br>(n=32) | Control Group<br>(n=29) | p value |
| --- | --- | --- | --- |
| **In hospital outcomes** |  |  |  |
| PPC (%) |  |  |  |
| No | 29 (90.6) | 24 (82.8) | 0.460 |
| Yes | 3 (9.4) | 5 (17.2) |  |
| HDU stay (%) |  |  |  |
| No | 31 (96.9) | 27 (93.1) | 0.600 |
| Yes | 1 (3.1) | 2 (6.9) |  |
| ITU stay (%) |  |  |  |
| No | 31 (96.9) | 28 (96.6) | 1.000 |
| Yes | 1 (3.1) | 1 (3.4) |  |
| Death (%) |  |  |  |
| No | 32 (100) | 29 (100) | N/A |
| Yes | 0 | 0 |  |
| Hospital LOS (days), median (IQR) | 4 (3-6) | 4 (3-6.5) | 0.336 |
| **Outcomes at 3 months** |  |  |  |
| BMI kg/m<sup>2</sup>, mean (SD) | 33.7 (4.13) | 35.8 (4.52) | 0.631 |
| Unplanned weight loss in past 3-6 months (%) |  |  |  |
| < 5% | 24 (80) | 16 (55.2) | **0.026** |
| 5-10% | 6 (20) | 8 (27.9) |  |
| ≥ 10% | 0 | 5 (17.2) |  |
| Missing | 2 |  |  |
| MUST Score 3 months (%) |  |  |  |
| 0 | 21 (70) | 17 (58.6) | 0.228 |
| 1 | 8 (26.7) | 6 (20.7) |  |
| 2 | 1 (3.3) | 5 (17.2) |  |
| 4 | 0 | 1 (3.4) |  |
| Missing | 2 |  |  |
| Wound complications (%) |  |  |  |
| No | 28 (90.3) | 27 (93.1) | 1.000 |
| Yes | 3 (9.7) | 2 (6.9) |  |
| Missing | 1 |  |  |
| Readmission (%) |  |  |  |
| No | 26 (81.3) | 23 (79.3) | 1.000 |
| Yes | 6 (18.8) | 6 (20.7) |  |
| Death (%) |  |  |  |
| No | 32 (100) | 29 (100) | N/A |
| Yes | 0 | 0 |  |

BMI, body mass index; MUST; Malnutrition Universal Screening Tool.

At 3-months there were significantly less patients who had unplanned weight loss in the intervention group compared to the control group (p=0.026); with 0 patients in the intervention group having 10% weight loss compared to 5 patients in the control group (17.2%) (**table 3**). There were no differences seen in wound complications and readmission between groups.

### Physiological outcomes

There were no detected differences between hand grip strength and peak expiratory flow rate between groups **(table 4)**.

**Table 4:** Physiological parameters

|  | Nutrition Group | Control Group | p value |
| --- | --- | --- | --- |
| Hand grip strength (kg/m<sup>2</sup>), median (IQR, n) |  |  |  |
| Baseline | 28.3 (23.3-36.2, 32) | 31.35 (23.9-34.3, 29) | 0.868 |
| POD1 | 28.3 (20-39, 27) | 27.95 (23.5-35.8, 22) | 0.972 |
| POD2 | 24.3 (20.6-36.2, 25) | 28 (18.9-35.2, 19) | 1.000 |
| POD3 | 23.9 (20.3-35.8, 23) | 29.3 (22.4-33.2, 14) | 0.546 |
| Discharge | 25.3 (20.6-37.3, 29) | 30.7 (22.4-35.7, 24) | 0.648 |
| Peak flow (l/min) mean (SD, n) |  |  |  |
| Baseline | 361.8 (160.4, 32) | 347.3 (112.2, 26) | 0.687 |
| POD1 | 182.3 (94.6, 26) | 192.8 (82.0, 18) | 0.698 |
| POD2 | 184.1 (89.3, 22) | 193.5 (88.5, 17) | 0.744 |
| POD3 | 168.1 (52.1, 21) | 182.1 (67.6, 14) | 0.517 |
| Discharge | 200 (85.0, 28) | 221 (90.7, 24) | 0.396 |

### Patient reported outcomes

*Results* from the questionnaire responses had uncertainty, and as with the clinical outcomes, the study was not powered to reach a statistical significant between groups (**table 5**). There was a trend for lower overall visual analogue scores in the nutritional intervention group at all timepoints, where patients with lower scores were less symptomatic. With regards to specific symptoms, at 3 weeks after surgery, in the intervention group there was significantly less thirst (2.6 vs 4.8; p=0.003) and mouth dryness (2.8 vs. 4.8; p=0.010), and a trend for reduced hunger (2.5 vs 3.7; p=0.055), but this did not reach significance. The median total QoR-40 score showed trend for higher in the interventional group at all timepoints other than baseline. The EQ-5D-5L Thermometer showed a trend for higher scores at all times points in the intervention group; higher scores in both of these represent better health. The EQ-5D-5L Scores showed trend for lower scores at all timepoints apart from baseline, where lower scores represent better health.

**Table 5:** Results of patient reported outcomes

|  | Nutrition Group<br>(SD, n) | Control Group<br>(SD, n) | Difference between<br>groups (95% CI) | p value |
| --- | --- | --- | --- | --- |
| *Visual Analogue Score* |  |  |  |  |
| *Thirst (0-10, higher = worse)* |  |  |  |  |
| Baseline | 4.0 (3.2, 32) | 4.9 (3.2, 29) | -0.8 (-2.5 to 0.8) | 0.314 |
| POD1 | 7.0 (3.6, 28) | 7.2 (3.4, 27) | -0.1 (-2.0 to 1.7) | 0.875 |
| POD2 | 5.6 (3.2, 26) | 6.2 (3.0, 24) | -0.6 (-2.4 to 1.2) | 0.506 |
| POD3 | 4.1 (3.4, 22) | 6.2 (2.8, 17) | -2.1 (-4.1 to -0.1) | 0.042 |
| Discharge | 4.1 (2.8, 30) | 5.3 (3.1, 28) | -1.2 (-2.7 to 0.4) | 0.146 |
| 3 weeks | 2.6 (2.7, 28) | 4.8 (2.5, 25) | -2.2 (-3.6 to -0.8) | **0.003** |
| 3 months | 2.5 (2.4, 28) | 3.0 (3.0, 26) | -0.4 (-1.9 to 1.0) | 0.564 |
| *Hunger (0-10, higher = worse)* |  |  |  |  |
| Baseline | 3.4 (2.5, 32) | 3.0 (2.5, 29) | 0.4 (-0.8 to 1.7) | 0.494 |
| POD1 | 2.4 (2.9, 28) | 3.6 (3.7, 27) | -1.2 (-2.9 to 0.7) | 0.207 |
| POD2 | 2.6 (2.6, 26) | 2.5 (2.6, 24) | 0.1 (-1.4 to 1.5) | 0.920 |
| POD3 | 1.5 (1.5, 22) | 2.1 (2.2, 17) | -0.7 (-1.9 to 0.6) | 0.295 |
| Discharge | 2.9 (3.0, 30) | 2.7 (2.1, 28) | 0.2 (-1.2 to 1.5) | 0.784 |
| 3 weeks | 2.5 (1.9, 28) | 3.7 (2.3, 25) | -1.1 (-2.3 to 0.03) | 0.055 |
| 3 months | 2.5 (2.1, 28) | 2.4 (2.5, 26) | 0.1 (-1.2 to 1.4) | 0.860 |
| *Mouth dryness (0-10, higher = worse)* |  |  |  |  |
| Baseline | 3.3 (2.7) 32 | 4.6 (3.8, 29) | -1.3 (-3.0 to 0.5) | 0.144 |
| POD1 | 8.4 (2.7) 28 | 7.6 (3.7, 27) | 0.8 (-1.0 to 2.6) | 0.368 |
| POD2 | 6.2 (3.3) 26 | 6.6 (3.2, 24) | -0.4 (-2.3 to 1.5) | 0.673 |
| POD3 | 4.2 (4.0) 22 | 5.5 (3.8, 17) | -1.2 (-3.8 to 1.3) | 0.329 |
| Discharge | 4.6 (3.1) 30 | 5.2 (3.7, 28) | -0.5 (-2.4 to 1.3) | 0.550 |
| 3 weeks | 2.8 (2.9) 28 | 4.8 (2.5, 25) | -2.0 (-3.5 to -0.5) | **0.010** |
| 3 months | 2.2 (2.4) 28 | 4.2 (3.8, 26) | -1.9 (-3.6 to -0.1) | **0.039** |
| *Weakness (0-10, higher = worse)* |  |  |  |  |
| Baseline | 1.8 (2.4, 32) | 1.6 (2.4, 29) | 0.2 (-1.1 to 1.4) | 0.797 |
| POD1 | 6.2 (3.2, 28) | 6.5 (3.1, 27) | -0.3 (-2.0 to 1.4) | 0.755 |
| POD2 | 5.1 (3.4, 26) | 6.0 (3.4, 24) | -0.9 (-2.9 to 1.0) | 0.346 |
| POD3 | 4.9 (2.9, 22) | 5.7 (3.5, 17) | -1.0 (-3.2 to 1.1) | 0.342 |
| Discharge | 3.9 (2.7, 30) | 4.4 (3.0, 28) | -0.5 (-2.0 to 1.0) | 0.490 |
| 3 weeks | 3.8 (3.0, 28) | 5.1 (2.3, 25) | -1.4 (-2.8 to 0.1) | 0.064 |
| 3 months | 2.7 (2.6, 28) | 3.4 (3.3, 26) | -0.7 (-2.3 to 0.9) | 0.389 |
| *Total (0-50, higher = worse)* |  |  |  |  |
| Baseline | 12.6 (8.2, 32) | 14.1 (8.1, 29) | -1.5 (-5.7 to 2.7) | 0.476 |
| POD1 | 24.1 (8.7, 28) | 24.9 (9.1, 27) | -0.8 (-5.6 to 4) | 0.745 |
| POD2 | 19.5 (7.8, 26) | 21.4 (7.6, 24) | -1.8 (-6.2 to 2.6) | 0.405 |
| POD3 | 14.5 (8.5, 22) | 19.5 (7.7, 17) | -5.0 (-10.3 to 0.3) | 0.062 |
| Discharge | 15.5 (8.5, 30) | 17.5 (8.4, 28) | -2.0 (-6.5 to 2.4) | 0.362 |
| 3 weeks | 11.7 (8.6, 28) | 18.4 (5.6, 25) | -6.7 (-10.7 to -2.7) | **0.001** |
| 3 months | 10.0 (7.9, 28) | 12.3 (10.5, 26) | -2.9 (-8.0 to 2.2) | 0.258 |
| *Patient Survey QoR-40 Total (40-200, higher = better)* |  |  |  |  |
| Baseline | 179.5 (15.5, 32) | 183.1 (9.9, 29) | -3.6 (-10.2 to 3.0) | 0.283 |
| POD1 | 162.7 (18.9, 28) | 159.1 (26.8, 27) | 3.6 (-9.0 to 16.2) | 0.568 |
| POD2 | 165.7 (17.3, 26) | 163.5 (18.7, 24) | 2.2 (-8.0 to 12.5) | 0.664 |
| POD3 | 163.9 (19.2, 21) | 157.6 (20.2, 17) | 6.3 (-6.8 to 19.4) | 0.337 |
| Discharge | 173.4 (16.1, 30) | 171.7 (21.2, 28) | 1.8 (-8.2 to 11.7) | 0.726 |
| 3 weeks | 174.9 (14.8, 28) | 168.1 (14.3, 25) | 6.8 (-1.2 to 14.8) | 0.095 |
| 3 months | 180.8 (15.8, 28) | 174.2 (20.8, 26) | 6.6 (-3.6 to 16.8) | 0.198 |
| *EQ-5D-5L Thermometer*<br>*(0-100, higher = better)* |  |  |  |  |
| Baseline | 78.5 (15.6, 32) | 75.1 (17.0, 29) | 3.4 (-5.0 to 11.8) | 0.421 |
| Discharge | 67.1 (18.4, 30) | 59.1 (22.1, 28) | 8.0 (-2.7 to 18.7) | 0.141 |
| 3 weeks | 70.0 (15.7, 28) | 63.8 (15.4, 25) | 6.2 (-2.4 to 14.8) | 0.151 |
| 3 months | 75.1 (23.5, 28) | 70.5 (21.7, 26) | 4.6 (-7.7 to 17.0) | 0.457 |
| *EQ-5D-5L Score*<br>*(0-25, higher = worse)* |  |  |  |  |
| Baseline | 7.1 (2.2, 32) | 6.9 (2.6, 29) | 0.1 (-1.1 to 1.4) | 0.833 |
| Discharge | 10.1 (2.2, 30) | 10.9 (3.3, 28) | -0.8 (-2.2 to 0.7) | 0.308 |
| 3 weeks | 8.7 (2.6, 28) | 9.7 (3.1, 25) | -0.9 (-2.5 to 0.7) | 0.263 |
| 3 months | 8.3 (3.0, 28) | 8.8 (4.0, 26) | -0.5 (-2.4 to 1.5) | 0.618 |

POD, postoperative day.

### Safety

There were no safety concerns expressed by the trial management group who met during the recruitment period. There were no deaths of patients during the study period. There were no serious adverse events recorded during the study period. The nutritional drinks were considered safe as in previous trials and there were no concerns during the study. None of the adverse events reported were deemed to be related to the study intervention.

### Qualitative interviews

Semi-structured qualitative interviews were undertaken in 14 patients randomised into the study. The interviews were conducted at 3-4 weeks post hospital discharge, the overall aim of the interviews was to ascertain if the trial processes were acceptable to the participants and to aid insight into trial intervention and questionnaires used to captures symptoms and recovery. The themes that emerged from the interviews around the trial consent, randomisation and impact on the hospital stay were positive, all patients felt well informed of the trial processes such as consent and randomisation. All participants reported that the study questionnaires captured recovery and general health and wellbeing throughout the surgical recovery and did not find the questionnaire burdensome.

## DISCUSSION

Our key indicators for feasibility were met. We have shown that a large multicentre RCT of nutritional intervention in major lung surgery with an objective of assessing postoperative outcomes is feasible. The study completed recruitment 5 months ahead of target. It is also possible to both randomise and follow up patients with high fidelity over the 3-month period. Importantly, qualitative interviews demonstrated that the trial design and the nutritional intervention were acceptable. Patients felt the questionnaires used captured their experience of recovery and symptom burden from surgery well. Patients who were randomised to the nutritional intervention group had less PPC incidence, although not powered to reach statistical significance. Unintentional weight loss was less in the nutritional intervention group at 3 months. The study questionnaires showed trends for increased perceived health in participants in the interventional group. There were significant differences found in symptom scores at 3 weeks, however this would need to be investigated further in a much more substantive trial.

This feasibility study has allowed the fine tuning of processes ahead of a larger more substantive trial. There would be modifications to the future study protocol. With regards to inclusion criteria, patients were included if they were having major lung surgery. Over 90% of our patients were having thoracic surgery for cancer and very few patients were excluded on the basis of the current eligibility criteria. As major lung surgery varies from biopsy to pneumonectomy, and some patients undergoing surgery may be having the procedure for other reasons than cancer, such as lung volume reduction surgery in COPD for example. We believe going forward the substantive study would focus on patients with newly diagnosed lung cancer who are having major thoracic surgery for lung resection. Therefore, the future large-scale study would include patients undergoing curative lung cancer surgery. Given the success of the recruitment process in this feasibility study, we feel this would not impact on recruitment rates of eligible participants. With regards to the exclusion criteria, we excluded patients with a BMI <18.5. The reason this was initially chosen as an exclusion criterion was because guidelines require those patients with BMI < 18.5 kg/m^2^ to have additional nutritional support in the form of ONS.^19^ The substantive study would need to incorporate these patients and recognise that patients may have different baseline nutritional needs in major lung surgery.^36^ Benefits may be more marked in patients with pre-existing malnutrition.^17,37^ Therefore, those with a BMI < 18.5 kg/m^2^ would be included, and participants could be randomised with aim to best balance patients with BMI < 18.5 or > 18.5 kg/m^2^.

The patients in the feasibility study were well balanced according to type of surgical approach (open or VATS). There were however significant differences in pack year history, with a higher number of never smokers in the intervention group. Smoking is the biggest risk factor for development of PPC and reduces following smoking cessation.^2^ A tailored smoking cessation intervention in the thoracic surgical pathway is currently being investigated in a feasibility study,^38^ though the optimum timing to stop before surgery is yet to be determined.^39^ The substantive study should therefore factor in smoking history into the randomisation component of trial design, with aim to balance current and recent quitters from long-term ex-smokers and never smokers.

With regards to the control, in this study a placebo was not required as the one or the outcomes of the study was whether the patients had the nutritional drinks or not. Therefore, there was no additional value in having placebo control for this reason. This study showed that patients are compliant with carbohydrate (CHO) loading and oral nutritional supplement (ONS) drinks with compliance of 89% with postoperative drinks and 100% of patients had at least 5 CHO loading drinks pre-operatively. There is no set definition for adherence to nutritional supplements however the results from this study replicate other research in this area. A systematic review into compliance of oral nutritional supplements in a range of settings and clinical conditions found mean compliance of 78%, ranging from 67% in hospital and 81% in community.^40^ Within cancer prehabilitation, compliance of 93.7% was demonstrated with whey protein supplement drinks^41^ and 100% compliance with carbohydrate loading.^42^ The substantive study would look to randomise to either nutritional intervention or usual care as the control, rather than use equal measures of water. A lack of blinding could potentially lead to bias regarding outcome measures. Thus, the importance of having both clinical outcomes and patient reported outcomes in the study. In our feasibility study both patient reported outcomes and clinical outcomes detected differences between groups, despite the study not being powered to do so.

Whilst the feasibility study was not powered to show a significant difference in PPC incidence, It is important for the substantive study to be able to determine if nutritional intervention has an impact on PPCs. Even modest improvements in the PPC rate would have massive cost savings through reduction ITU admissions, hospital bed days used and readmission rates. In the UK 30-day readmission to hospital after lung cancer surgery is high (12%), and is a key target for improvement in the national lung cancer audit.^43,44^ Thus, potential clinical and cost benefits to the NHS of optimising nutrition and so ameliorating major complications and hospital readmission is significant.

There is growing evidence around the implementation of prehabilitation within surgical pathways. A systematic review found that multimodal prehabilitation involving exercise and nutrition has a positive impact on physical function in patients awaiting lung cancer surgery.^45^ The benefits of prehabilitation include personal empowerment, physical resilience and improvements in long term health.^46^ The substantive study could therefore embed the nutritional intervention as part of an enhanced package of multimodal prehabilitation compared to standard care, and the impact on postoperative outcomes, physical status and quality of life.

Since the study was not a definitive trial, the findings must not be overinterpreted. We cannot expect at this stage for the outcomes of the study to influence clinical care, as the study was not large enough to detect realistic sized differences in rates of post-operative outcomes. Also having only included one centre was not wide enough in terms of the number of centres involved in the feasibility study to reach a generalised result. However, this feasibility study is an important precursor to the larger, substantive trial and provides important information that will help ensure the success. A definitive study is needed to determine the impact of nutritional supplementation in thoracic surgery on both patient reported outcomes and postoperative outcomes. The full randomised controlled trial will allow the definitive answer to this question.

## Data Availability

All available study data can be obtained from the corresponding author.

## Acknowledgements

The thoracic research team at University Hospitals Birmingham NHS Foundation Trust for their support of the study.

## Contributors

BN and AK conceived the study. BN, AK, AS and NE were involved in trial design. AK was involved in patients’ recruitment and data acquisition. HF, RS, EB, MK and BN were involved in the surgical care of the participants. AK and SK were involved in patient interviews. AK, STL, AS and NE were involved in data analysis. AK and STL drafted the final manuscript. KK provided input from a dietetics perspective. All authors critically appraised and approved the final manuscript before publication.

## Disclaimer

The funder had no role in study design data collection, interpretation or analysis in writing the final manuscript for publication.

## Patient consent for publication

Not required.

## Ethics approval

Ethical approval was granted from Wales REC 7 research ethical committee REC reference 16/WA/0254, followed by HRA approval. The study was then registered with ISRCTN number: 16535341.

## Data sharing statement

All available study data can be obtained from the corresponding author.

